# SEN support from the start of school and its impact on unplanned hospital utilisation in children with cleft lip and palate: a demonstration target trial emulation protocol using ECHILD

**DOI:** 10.1101/2022.04.01.22273280

**Authors:** Vincent Grigori Nguyen, Anna Zylbergsztejn, Katie Harron, Tamsin Ford, Kristine Black-Hawkins, Kate Boddy, Johnny Downs, Martin Doyle, Matthew Lilliman, Jacob Matthews, Stuart Logan, Jugnoo Rahi, Ruth Gilbert, Lorraine Dearden, Bianca De Stavola

## Abstract

Special Educational Needs (SEN) provision for school children provides extra support and reasonable adjustments for children and young people with additional educational, behavioural or health needs to ensure equal education opportunities; for example those born with a healthcare need such as cleft lip and palate may be provided SEN to aid with challenges in communications. However, there is limited knowledge of whether SEN provisions impact academic or health outcomes in such a population and conducting a randomised controlled trial to establish this evidence is not plausible. In lieu of randomised controlled trials, target trial emulation methods can be used in attempt to answer causal questions using observational data whilst reducing confounding and other biases likely to arise with such data. The Education and Child Health Insights from Linked Data (ECHILD) dataset could be used as part of trial emulation methods to understand the impact of SEN provisions on academic and healthcare outcomes. ECHILD is the first dataset to hold longitudinal school, health and social care data on all pupils in England, obtained by linking the National Pupil Database (NPD) with Hospital Episode Statistics (HES). In this protocol, we describe how the ECHILD dataset could be used to explore and conduct a target trial emulation to evaluate whether children who were born with cleft lip and palate would have different unplanned hospital utilisation if they received or did not receive SEN provisions by Year 1 (specifically by January in their second year of school) when they are aged 5 or 6.

**Methods:** Focussing on the <u>population</u> of children who are identified as having been born with cleft lip and palate, an <u>intervention</u> of varying levels of SEN provision (including no SEN provision) by January of the second year of school, and an <u>outcome</u> of unplanned hospital utilisation, we apply a trial emulation design to reduce confounding when using observational data to investigate the causal impact of SEN on unplanned hospital admissions. Our target population is children born 2001-2014 who had a recording of cleft lip and palate in HES and who started their second year of primary school (Year 1) in a state school between 2006 and 2019; children with a first recording of cleft lip and palate after Year 1 were excluded (these were pupils who likely immigrated to England after birth). We intend to use a time window of SEN provision assignment between the start of school (reception) and by the January school census in Year 1. Using target trial emulation, we aim to estimate the average treatment effect of SEN provision on the number of unplanned hospital admissions (including admissions to accident and emergency) between the January school census in Year 1 and Year 6 (the end of primary school, when children are 10-11 years old).

**Ethics and dissemination:** Permissions to use linked, de-identified data from Hospital Episode Statistics and the National Public Database were granted by DfE (DR200604.02B) and NHS Digital (DARS-NIC-381972). Ethical approval for the ECHILD project was granted by the National Research Ethics Service (17/LO/1494), NHS Health Research Authority Research Ethics Committee (20/EE/0180) and UCL Great Ormond Street Institute of Child Health’s Joint Research and Development Office (20PE06). Stakeholders (academics, clinicians, educators and child/young people advocacy groups) will consistently be consulted to refine populations, interventions and outcomes of studies that use the ECHILD dataset to conduct target trial emulation. Scientific, lay and policy briefings will be produced to inform public health policy through partners in the Department of Education and the Department of Health and Social Care.

## Introduction

Special Educational Need (SEN) provisions aim to deliver reasonable adjustments in children and young people (CYP) in a school environment who need additional health, educational, or behavioural support, including children with complex health needs or learning disability. SEN provides support to those in need using a variety of facilities including support from a teaching assistant, assistance with communication, special learning programmes and support with physical needs. SEN provisions are divided into two levels, including: SEN Support (this is often known as Action, Action Plus or non-Statemented SEN) and Education and Health Care Plan (EHCP, this is previously known as Statemented SEN). SEN Support is organised at a school or college level and provides access to children and young people in need of SEN with support that may include teaching assistants who aid in communications, special learning programmes and supporting physical needs. An EHCP is organised by local authorities for children and young people who require further adjustments and often require additional resources (compared to SEN Support) to aid in education, health and social care needs.

Currently, there is limited research on the impact of SEN on academic and healthcare outcomes in populations who are in need of SEN. To establish the causal effect of SEN on outcomes, randomised controlled trials would have to be conducted, however such study designs are not always feasible due to the human, time, financial and ethical costs associated. As SEN support is universally available in primary schools in England, conducting a randomised controlled trial would be unfeasible and possibly unethical at least for certain groups of children. In lieu of randomised controlled trials, observational studies provides a pragmatic, data-driven, observational study alternative when trials are not possible. One major challenge with using observational data when compared to data collected from randomised controlled trials is the risk of confounding, particularly, confounding by indication where assignment to treatment is not random and is often related to the severity of a medical condition. However, attentive study design can mitigate such biases in observational data by emulating the protocol of an equivalent randomised controlled trial (Hernán and Robins, 2016).

An example of dataset that can used to evaluate the causal effect of SEN on healthcare outcomes using trial emulation is the ECHILD dataset, (https://www.adruk.org/our-work/browse-all-projects/echild-linking-childrens-health-and-education-data-for-england-142/). The ECHILD dataset is the first dataset in England to link academic data with secondary care hospital data for all pupils, and can be used to investigate the associations between health, education and social care (Mc Grath-Lone et al., 2021). The ECHILD dataset links data from the National Pupil Database (NPD) and Hospital Episode Statistics (HES) and currently includes children and young people in England who were born between 1 September 1995 and 31 August 2020. Therefore, the ECHILD dataset provides the opportunity to conduct observational studies with long-term follow-up; currently follow-up is up to age 25 years (from birth in 1995 until hospitalisations in 2020). With data on clinical conditions, social care status, hospital visits, academic attainment and SEN provisions in school, the ECHILD dataset enables investigation of associations between a variety of exposures and outcomes when adjusting for different confounders in populations at risk. Examples of population at risk that can be phenotyped in hospital data include children with major congenital anomalies (e.g. Down Syndrome), cerebral palsy, learning disabilities, epilepsy, diabetes, and premature birth. Exposures in the education data include provision of support for SEN, Free School Meal status and measures of deprivation. Outcomes in ECHILD potentially include both educational and health related outcomes such as (specific types of) hospitalisations, national examination results (at multiple key stages) and exclusions from school.

In this study protocol, we describe how we aim to use the ECHILD dataset and design an appropriate study that emulates a target trial that would address the question of interest. As an initial demonstration of the ECHILD dataset, we focus on the impact of receiving different types of SEN provision (including none) on unplanned hospital utilisation in children who were born with cleft lip and palate (Bell et al., 2016). We address how to evaluate whether an intervention recorded in educational data can impact outcomes recorded in health data.

Incidences of cleft lip and palate impact 900 newborns in England yearly and impact communications (hearing and speech), dental health (Gallagher and Collett, 2019) and psychosocial health. Cleft lip and palate are associated with lower academic attainment (Fitzsimons et al., 2018) and have been linked to a three-fold increase in hospitalisations in Australia when compared to those without cleft lip and palate for all ages (Bell et al., 2016). Prior observational studies have suggested that extra support when starting school may be beneficial to children with cleft lip and palate for academic outcomes (Fitzsimons et al., 2018). Whilst Fitzsimons et al., 2018, has shown that children with cleft lip and palate are more likely to receive SEN provisions, particularly the types of SEN that concentrate on speech, language and communication support, to our knowledge, no study has compared the impact of extra help in terms of SEN using a control group of children with cleft lip and palate who have less or no support for any outcome.

## Methods

### Study Design and Setting

The study will be a retrospective observational study based on data from the ECHILD dataset. The ECHILD database has been described previously (Mc Grath-Lone et al., 2021). Briefly, the ECHILD dataset consists of pseudo-anonymised linked data from the National Pupil Dataset and Hospital Episode Statistics for children who were born between 1 September 1995 and 31 August 2020 in England.

### Dataset and Linkage

The data source we will use is the ECHILD database, a pseudo-anonymised dataset that links the NPD with HES. A full description of the ECHILD dataset has previously been described (Mc Grath-Lone et al., 2021). In brief, the ECHILD’s extract of the National Pupil Database contains data from academic terms (Summer, Autumn and Spring) between 2006 and 2020 and contains details on school, local authority, age, gender, ethnicity, first language, socioeconomic status, free school meal status, absence related data, social care/children in need related data and SEN status. The National Pupil Database is considered near universal as it covers all state schools in England from Key Stage 1 to Key Stage 4.

The ECHILD’s extract of Hospital Episode Statistics contains details on admitted patient care, outpatient appointments, accident and emergency utilisation, and critical care between 1997 until 2021, It contains details on age, gender, ethnicity, clinical information recorded during hospital admissions, including details of diagnoses, and operations. Hospital Episode Statistics is considered near complete as it covers 99% of public hospital activity in England (Herbert et al., 2017). HES records since 1998 are also linked to ONS Mortality data covering information on causes and timing of deaths. The linkage coverage periods are described in (Mc Grath-Lone et al., 2021) and ECHILD has been shown to have a high linkage rate between NPD and HES (Libuy et al., 2021a) of 95%. Such high linkage rates are attributable through a two-stage linkage process (Libuy et al., 2021b).

### Population

Our population consists of children born with a cleft lip and palate and followed from Year 1 of school (the second year of formal school attendance) between 2006 and 2019 (i.e. born between 2001 and 2014). To identify pupils who started Year 1 between 2006 and 2019, the earliest recording of “1” from the NCActualYear (National Curriculum Actual Year) variable in the NPD dataset will be used. To identify pupils with cleft lip and palate, ICD10 codes will be applied to HES diagnoses and OPCS codes will be applied to operations (Appendix 1 - Table 1 - Table 3); for each pupil, the earliest recorded date in HES would be considered the “diagnosis” date. We will remove pupils whose first recording of cleft lip and palate in HES is after their first year in school to evaluate the incident use of SEN; however as cleft lip and palate are often identified at or before birth, we suspect this would encompass a small number of pupils, for example, those born outside of England. To categorise cleft severity, we will use the following categories: cleft lip only, cleft palate only, unilateral cleft lip and palate, and bilateral cleft lip and palate(algorithm/method in Appendix 1 Table 2).

### Intervention and Exposure Variables

The intervention assessed is the level of SEN (including none) recorded by Year 1 of school (age 5 to 6); whilst SEN support status can change throughout a child/young person’s educational journey, our implementation of trial emulation focusses on an intention-to-treat analysis (ITT), that is it analyses the assignment of treatment and not whether treatment was consistently adhered to (or provided).

Due to the potential needs based on the severity of cleft lip and palate, we aim to classify the exposure variable as “levels of SEN” as opposed to a binary outcome (i.e. SEN vs no SEN).This is because in the event that the majority of Bilateral Cleft Lip + Palate (or more severe types of cleft lip and palate) contains small numbers in terms of “no SEN”, we could compare “SEN Support” against “EHCP”. For this reason, our exposure variable would be classified as: No SEN, SEN Support and EHCP by Year 1 and use the appropriate level as reference (i.e. SEN Support would be used as the “control” group if all children had received some form of SEN). To establish SEN status at Year 1, we will use the January (Autumn) census in Year 1 of school due to funding being calculated using these censuses. See Table 1 for a list of variables describing SEN in the NPD.

In Figure 1 we demonstrate how we will identify the population and each subpopulation (based upon cleft severity), and how they will be classified according to the exposure variables (levels of SEN).

**Table 1:** List of variables recording SEN in the National Pupil Database (NPD)

| Variable Name in NPD | Variable Description as per the NPD data dictionary |
| --- | --- |
| PrimarySENtype | Nature of pupil's primary special educational need. For pupils with a SEN status of E or K their main or primary need and, if appropriate, their secondary need, should be recorded. |
| SecondarySENtype | Nature of pupil's secondary special educational need. For pupils with a SEN status of E or K their main or primary need and, if appropriate, their secondary need, should be recorded. |
| SENProvision | Provision types under the SEN Code of Practice |
| CensusSEN | Provision types under the SEN Code of Practice. |
| SEN_provision | Special Educational Needs provision |
| SENA<br>SENELK<br>SENELSE<br>SENF<br>SENPS<br>SEN_ALL<br>SENAPK<br>SENSE | • SENA: Does a pupil have SEN - school action?<br>• SENELK: Does pupil have SEN support<br>• SENELSE: Does pupil have SEN with statement or EHC plan<br>• SENF: Pupil SEN status<br>• SENPS: Does pupil have SEN - Action Plus or Statemented?<br>• SEN_ALL: Does pupil have SEN with or without statement or EHC plan?<br>• SENAPK: Does pupil have SEN without statement or EHC plan<br>• SENSE: Does pupil have SEN with statement or EHC plan |
| LatestSEN | Provision types under the SEN Code of Practice. |
| SENProvisionMajor | Pupil's major SEN provision group based on SEN provision code. |
| SENstatus | Provision types under the SEN Code of Practice. |
| SENUnitIndicator | Indicates if a pupil with SEN in a mainstream school is a member of a SEN Unit (sometimes called special class) |
| SpecialProvisionIndicator | Indicates if a pupil with SEN in a mainstream school is a member of an SEN Unit, special class or resourced provision. |

**Figure 1:**
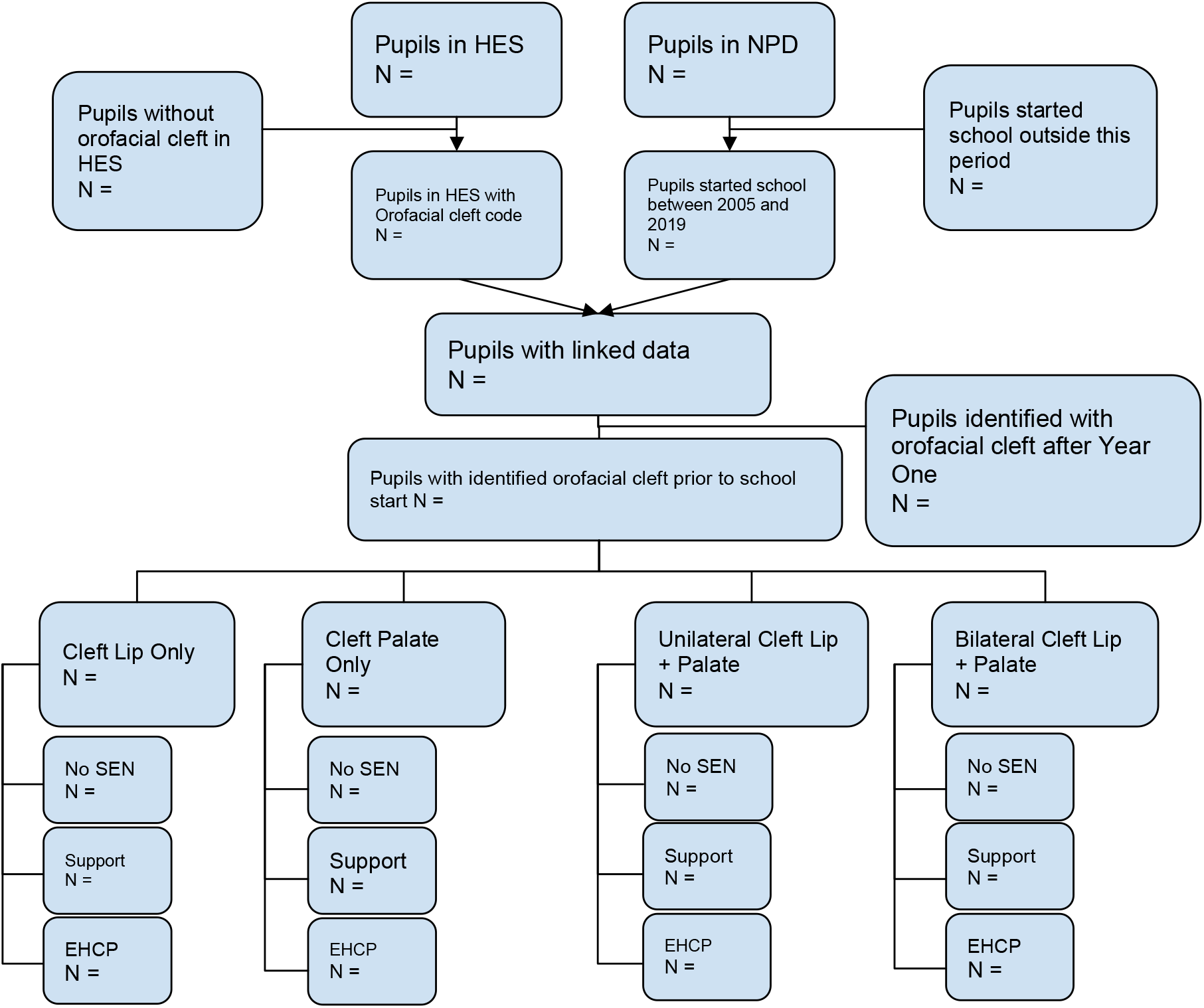
Flowchart of how the proposed population would be derived. Our population would consist of those who are identified in HES with cleft lip and palate before Year 1 and, who are linkable to NPD. To allow for heterogeneity of effects we aim to sub-classify our population based upon cleft lip and palate severity. SEN numbers will be based upon SEN recorded by Year 1. No SEN represents either never receiving SEN or receiving SEN after Year 1.

### Follow-up

Our population will be followed-up from the time exposure is assessed (January Census of Year 1) till the end of primary school (Year 6) or end of study whichever occurred first. Children and young people may be considered lost to follow-up if they were off rolled (no longer in education), transferred outside of an English school that received government funding or died (based upon linked Office of National Statistics mortality linkage).

Whilst the ECHILD data can be used to follow up children beyond the end of primary school (and for some individuals, beyond secondary school), we will limit our follow-up period in this protocol to the end of Year 6 for two reasons: 1) changing to secondary (or middle) school will re-evaluate the need for SEN, and many pupils may no longer be provided SEN, and 2) the time between the assignment of SEN considered here and the outcome may be too long if beyond Year 6, with the outcome affected by many intermediate factors (pupils starting Year 1 in 2006, would have 15 years of follow-up in HES).

### Outcome Variables

The primary outcome variable is unplanned hospital utilisation. To identify unplanned hospital admissions in Admitted Patient Care, we will use the admission method variable in HES (admimeth) (Appendix 2 for the case definition from HES Admitted Patient Care). For hospital utilisation that did not require an overnight admission, we will use the HES Accident and Emergency dataset to account for non-admitted unplanned hospital utilisation (Harron et al., 2018). We aim to combine Admitted Patient Care and “Accident and Emergency” datasets to create a timeline of unplanned hospitalisation between the Autumn census in Year 1 and the end of Year 6.

To measure unplanned hospitalisations, we aim to use two methods 1) the number of unplanned hospital admissions and 2) the number of unplanned days in hospital from study entry until study exit. When calculating the number of admissions using HES, we plan to combine admissions where the discharge date of one admission (disdate) was within one day of the next admission’s admission date (admidate).

### Covariates

To account for non-random SEN assignment to this population of children, we will use information on several covariates that are known or suspected to influence SEN assignment and the outcome of interest. These include socio-demographic, health-related and education-related variables referring to the child. Specifically, these included: gender, Income Deprivation Affecting Children Index (IDACI) quintiles for lower super output area of residence, free-school meal status, comorbidities, birth month, clinical treatments for cleft lip and palate prior to SEN level assignment, birth characteristics such as gestational age and birth weight and school and local authority level data. An additional school-level variable we aim to include is the type of school (mainstream or special). We will also include the interaction between type of school and level of SEN to account for the likely heterogeneity in the effect of SEN across schools.

The distribution of these potential confounders by population subgroup will be examined (see table 2 for an outline) and directed acyclic graphs representing the assumed relationships among these variables, SEN exposure, and the outcome of interest will be drawn to identify the variables that will be controlled for within the analysis using DAGitty ver 3.0.

**Table 2:**
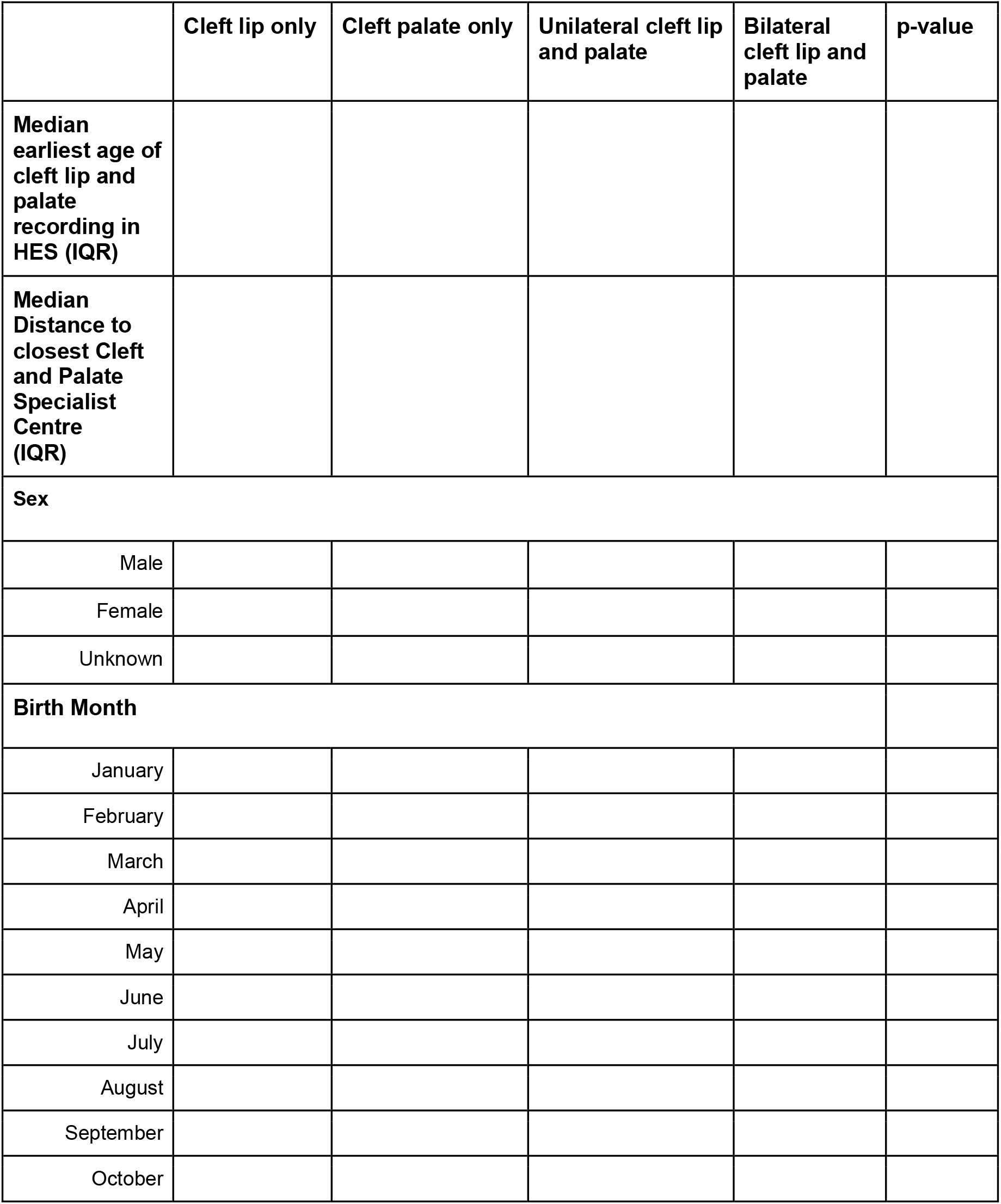
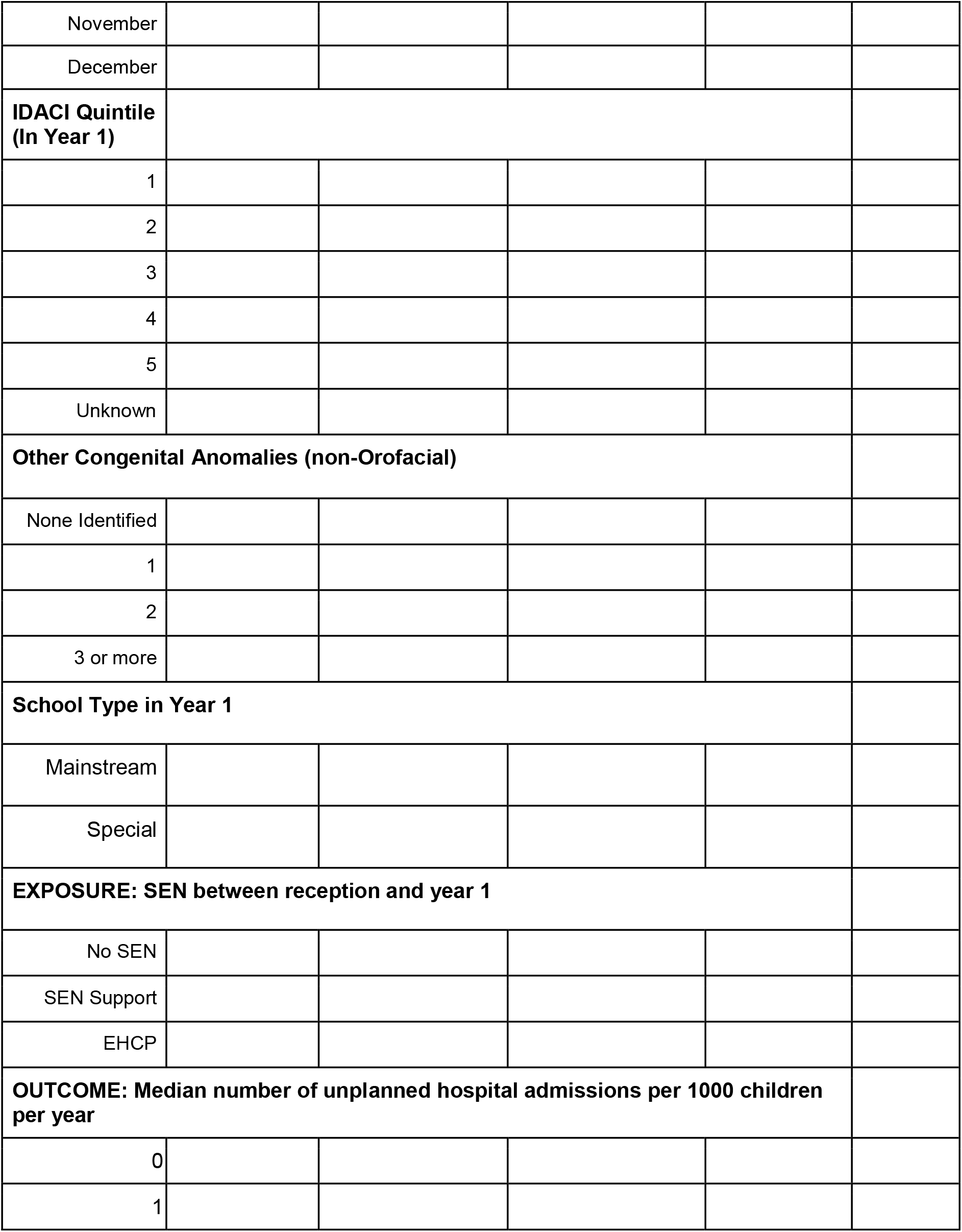
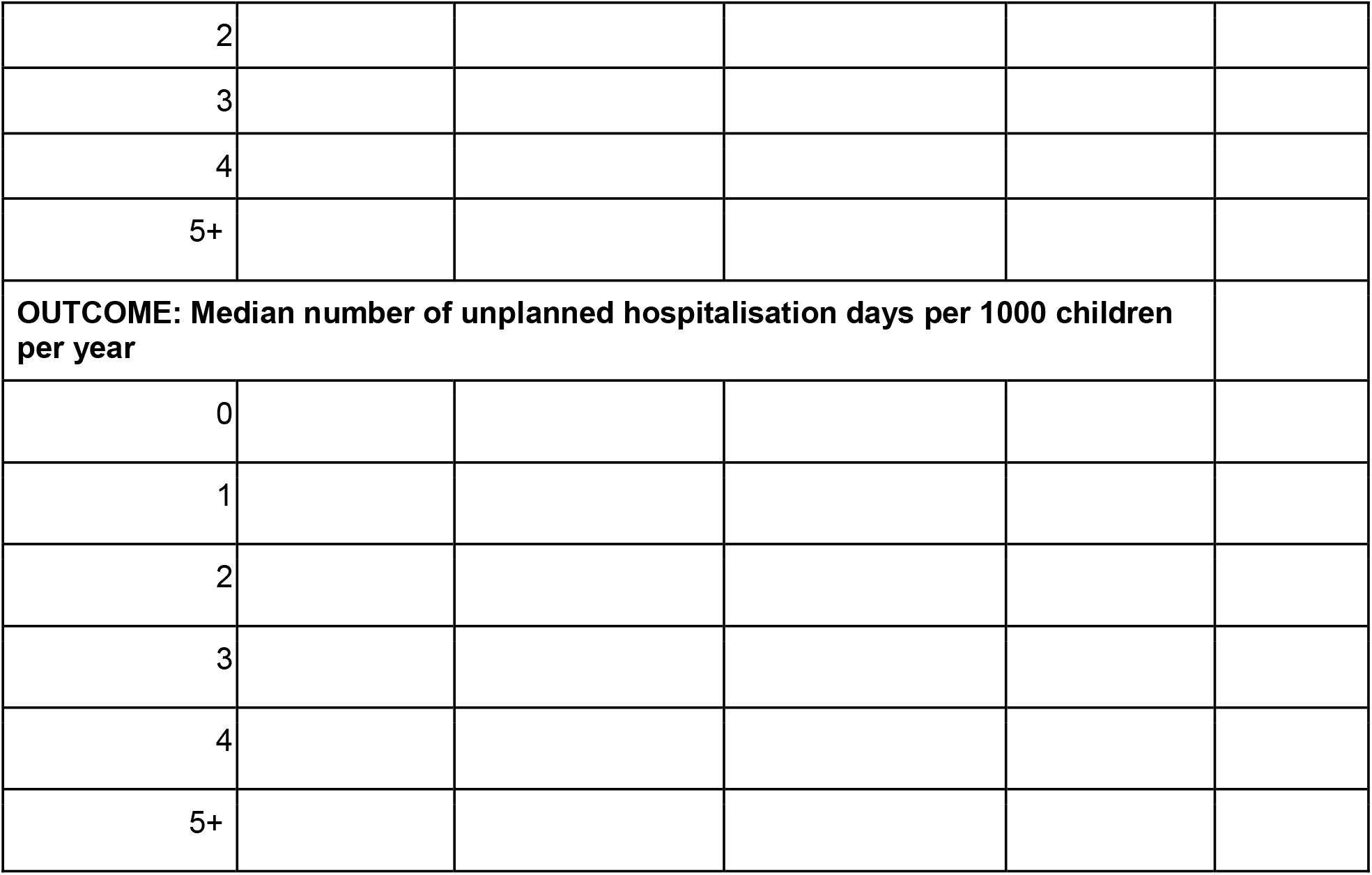
Demographics Table of the Distribution of the sociodemographic, educational and health characteristics amongst children with different severity of Cleft. The table will include N and row percentages (when applicable)

### Bias

To mitigate the likely confounding bias affecting observational data, we will adopt the Target Trial Emulation (TTE) framework. TTE enables observational data to be mapped to a hypothetical target experimental trial counterpart by creating the specification of an ideal (pragmatic) trial and using this as a basis to shape the observational study design. TTE consists of (1) defining the specifications of a hypothetical target experimental trial of the causal question of interest (including the corresponding effect), (2) emulating the specifications of the ideal target trial using observational data and (3) estimating the effects of interest using the emulated trial data. The first component of TTE involves defining an inclusion/exclusion criterion on entry, a treatment strategy (including time), follow-up frequency and modality, outcome measures, effects of interest (estimands) and the estimation method. Using the second component of TTE, observational data would be wrangled to emulate the distribution of the data if it were to have been gathered in the target trial. Finally, the third component of TTE requires the adjustment for confounding and is dependent on treatment assignment to aid in deciding the analytical methodology. In Table 3 we describe the target (ideal) trial one would design to investigate the causal effect of SEN provision (by January of the second year of school) in cleft lip and palate children and the equivalent emulated trial to be generated from ECHILD.

**Table 3:** Trial Emulation Framework to estimate the causal effect of SEN by Year 1 on unplanned hospitalisations in children with cleft lip and palate.

|  | **Ideal trial** | **Emulated trial** |
| --- | --- | --- |
| **Eligibility Criteria** | Geography: England<br>Started Year 1 in a state school between 2006 and 2019<br>Diagnosed with cleft lip and palate prior to Year 1 | Geography: England<br>Started Year 1 in a state school between 2006 and 2019<br>identified in HES with cleft lip and palate before start of year 1 |
| **Recruitment period** | 2006 to 2019 | 2006 to 2019 staggered by start year to account for structural changes through time (See Figure 2) |
| **Follow-up duration** | From: Start of SEN or placebo<br>To:<br>Until the end of primary school OR<br>Loss of follow-up (e.g., emigration) OR<br>Death OR<br>End of study | From: January Census in Year 1<br>To:<br>Until the end of primary school OR<br>Loss of follow-up in NPD OR<br>Death OR<br>End of study/end of data (HES outcomes until 30 August 2020) |
| **Outcome(s)** | Unplanned hospital Utilisation as defined by:<br>Counts of unplanned hospital admissions<br>Counts of unplanned hospitalisation days | Unplanned hospital Utilisation as defined by:<br>Counts of unplanned hospital admissions in HES<br>Counts of unplanned hospitalisation days in HES |
| **Treatments to be compared** | Level of SEN assigned between the start of reception and the end of Year 1<br>Lowest Level of SEN (including No SEN) assigned to be used as a control group between the start of reception and Year 1 | Level of SEN recorded by the January census in Year 1<br>Lowest Level of SEN (including No SEN) to be used as a control group recorded between the start of reception and Year 1 |
| **Estimand** | The average treatment effect of initiating SEN by Year 1 on the rate of unplanned hospital admissions expressed as rate ratios and rate differences<br>The average treatment effect of initiating SEN by Year 1 on the number of unplanned hospital days expressed as a rate ratios and rate differences | The average treatment effect of initiating SEN by Year 1 on the rate of unplanned hospital admissions expressed as a rate ratios and rate differences<br>The average treatment effect of initiating SEN by Year 1 on the number of unplanned hospital days expressed as a rate ratios and rate differences |
| **Analysis plan** | Poisson or Negative Binomial Regression (or zero-inflated regression, depending on the degree of overdispersion) of the number of events accountings for duration of follow-up.<br>Clustering by school and/or local authority to be dealt with using either mixed effects models or robust inference (e.g., GEE). | Appropriate methods for confounding adjustment (such as regression adjustment and standardisation, propensity score-based methods) involving Poisson or Negative Binomial Regression (or zero-inflated regression, depending on the degree of overdispersion) of the number of events accountings for duration of follow-up.<br>Clustering by school and/or local authority to be dealt with using either mixed effects models or robust inference (e.g., GEE). |

### Analysis

#### Explorative analyses

To explore confounding by indication in the causal relationship between receiving (different levels of) SEN and the outcome(s) of interest in the target population, possibly stratified by subtype, we will use the ECHILD dataset and conduct exploratory analysis to understand 1) whether the proportion and type of SEN provision differs between severity of cleft lip and palate and 2) whether such proportions change between 2006 and 2019, separately by category of cleft lip and palate. Such analysis would allow us to understand whether the severity of the cleft lip and palate is related to receiving SEN, whether certain severities of cleft lip and palate would have enough exposed/unexposed participants and whether public policy changes through time have impacted these proportions. Figures 2 and 3 are examples using simulated data of possible representations of how cross-sectional and cumulative frequencies of SEN provision and type vary over time, separated by cleft lip and palate type.

**Figure 2:**
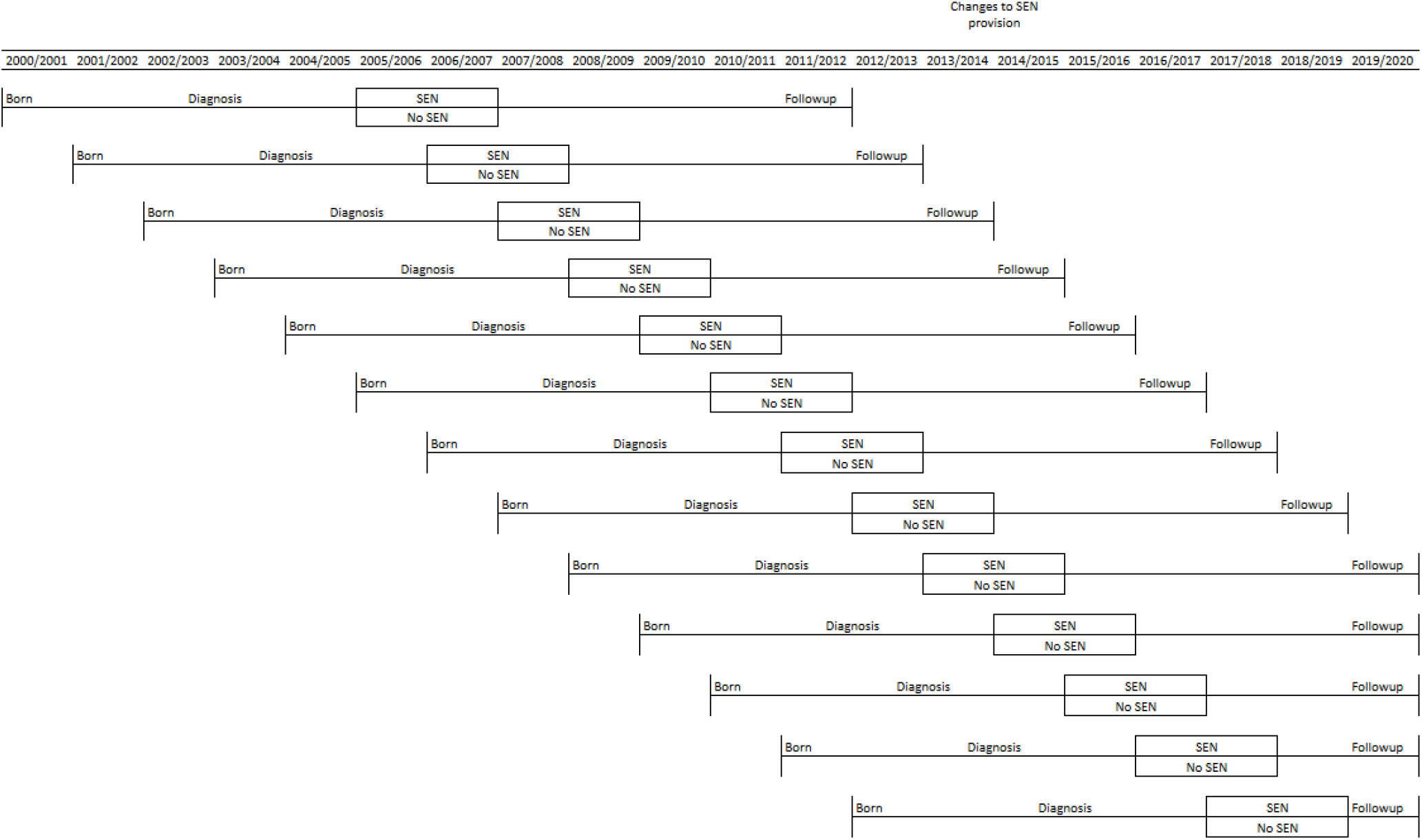
Example figure of how the single ECHILD cohort is segmented into multiple sequential cohorts including diagnosis period, exposure window and follow up period.

**Figure 2:**
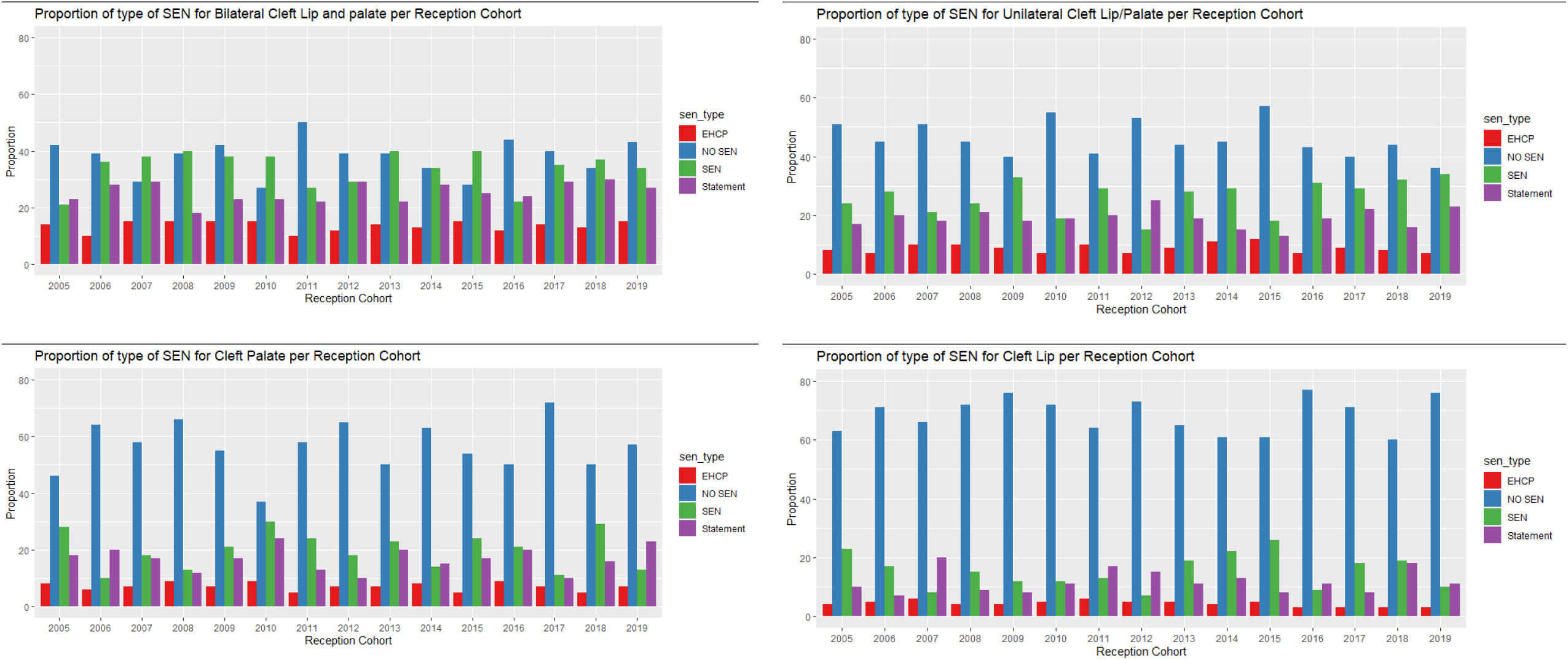
Simulated data showing different proportions of types of SEN per reception cohort, per cleft severity level

**Figure 3:**
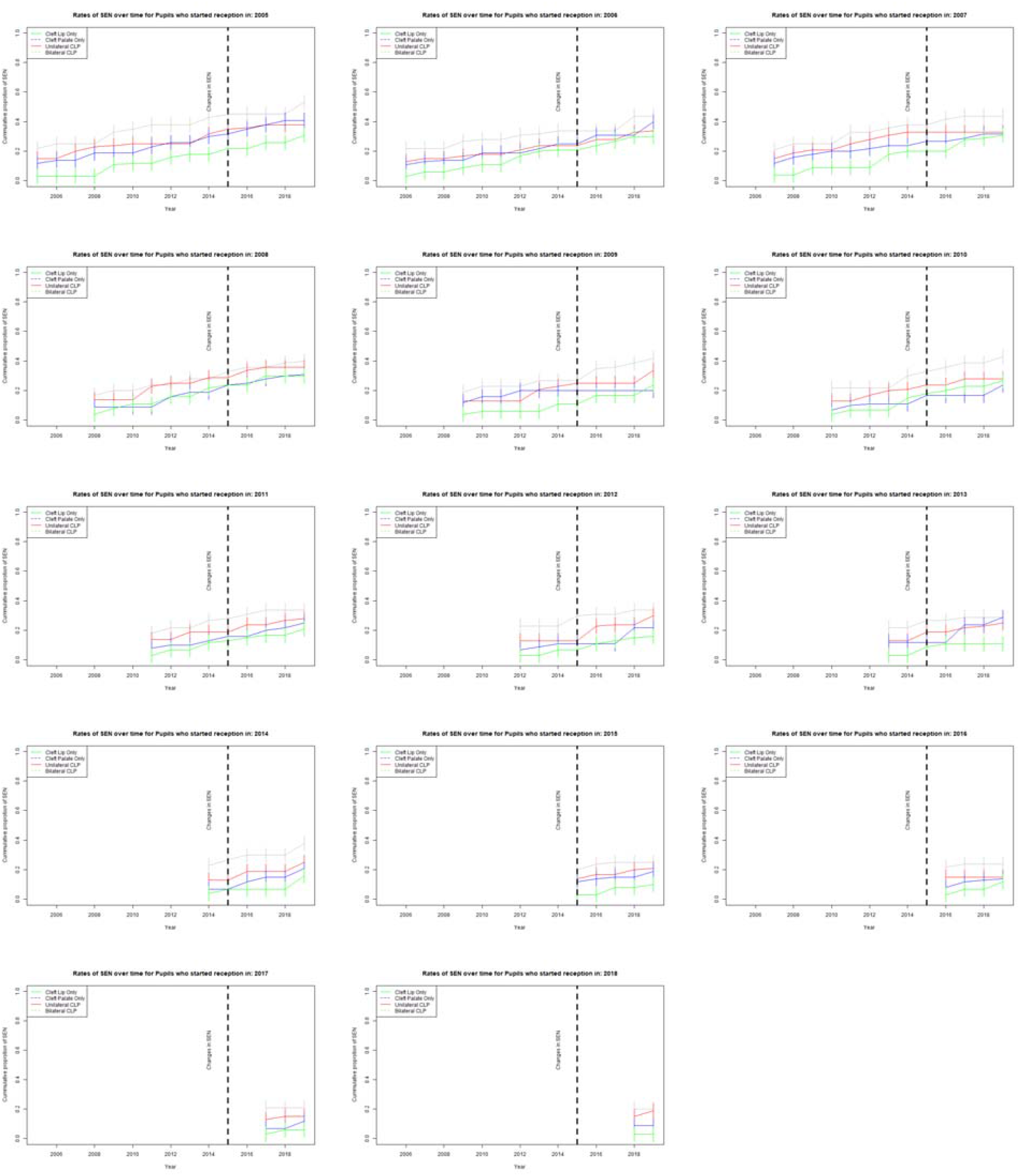
Simulated data showing the cumulative rates of SEN for each cohort and cleft severity over time

#### Causal analyses

Once explanatory analyses have been conducted, alternative estimation methods will be employed to address confounding bias due to non-random assignment to SEN. We will compare results obtained assuming no-unmeasured confounding (that is, we have data on all the relevant confounders) and assuming instead that we have an instrumental variable (if there is for example variation in provision by local authorities). The first group of methods will include: outcome-based models such as g-computation and propensity score methods (Inverse probability weighting and matching) using the covariates previously mentioned. The second group of methods will only be possible if suitable instruments for SEN provision are identified, for example if there are policy changes in provision that are implemented at different times across local authorities and will use instrumental variable-based estimators.

As we aim to model the number of admissions and the days spent in hospital, we will use Poisson (or negative binomial) outcome models with the logarithm of follow-up time (expressed as number of terms) as offset. The likely clustering of pupils within local authority and school will be addressed either by fitting mixed effects models or by using robust inference (or both).

#### Missing Data

Based upon the proportion of missingness in the data and the mechanisms of missingness, we will first use complementary non-missing data points in HES and NPD prior to data imputation; for example, using Sex variable from HES to complement missing values in the NPD variable Gender. We aim to use multiple imputation using chained equations to generate multiple possible (for example, ten alternative) values for each missing data point (Azur et al., 2011). In the event we use both multiple imputation and matching, we will conduct imputation first and then match in each alternative dataset (Leyrat et al., 2017)

#### Sensitivity Analyses

To account for uncertainty in the recording of observational data such as measurement errors, we aim to conduct sensitivity analyses. First, we will conduct a sensitivity analysis to mitigate against a delayed recording in SEN provisions, by expanding the exposure window to the first term in Year Two as part of the Autumn census.

Second, we will conduct sensitivity analyses to account for the complexity of congenital abnormalities in addition to cleft lip and palate, stratifying by (instead of adjusting for) congenital abnormalities in children and young people with cleft lip and palate. See Figure 4 for the alternative population derivation and Appendix 4 for the socio-economic, educational and healthcare distribution table for each cleft severity and birth anomaly interaction.

**Figure 4:**
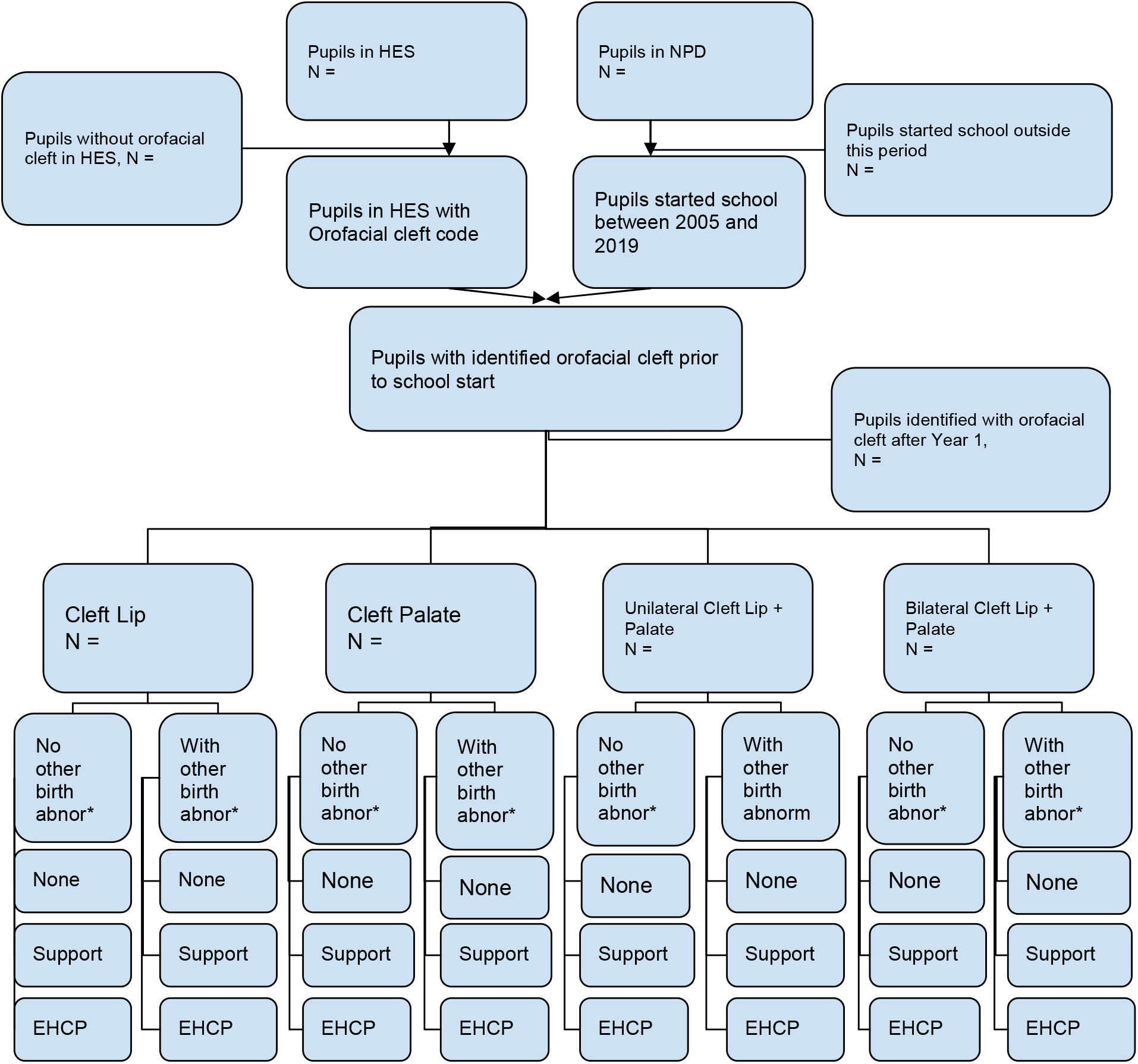
Alternative study design that stratifies for additional complexities in contrast to adjusting for comorbidities. Abnor* = abnormalities

## Stakeholder involvement

Prior to developing this protocol, two independent meetings were conducted with stakeholders (parents, pupils, teachers, etc) to understand which medical conditions are of interest and at which entry timepoints are important for child development. The first meeting was with the Department for Education’s national young SEN advisory group (FLARE) on the 18th of September 2021 and the second with the Young Persons Advisory Group for research at Great Ormond Street Hospital on the 27th November 2021. This engagement identified that school entry is an important key milestone when SEN provisions are required. Therefore, in the proposed study, we have used school start as our entry point and will generate further target trials based upon further patient engagement.

## Data Availability

Aggregate results from the ECHILD dataset will be pre-printed, revised and published. Individual record-level data with personal identifiers removed is currently hosted on the Office for National Statistics Secure Research Services data-sharing service. We are grateful to the Office for National Statistics (ONS) for providing the trusted research environment for the ECHILD Database. ONS agrees that the figures and descriptions of results in the attached document may be published. This does not imply ONS acceptance of the validity of the methods used to obtain these figures, or of any analysis of the results.
The ECHILD Database uses data from the Department for Education (DfE). The DfE does not accept responsibility for any inferences or conclusions derived by the authors. This work uses data provided by patients and collected by the National Health Service as part of their care and support. Source data can also be accessed by researchers by applying to NHS Digital.

## Ethics and dissemination

Permissions to use linked, de-identified data from Hospital Episode Statistics and the National Public Database were granted by DfE (DR200604.02B) and NHS Digital (DARS-NIC-381972). Ethical approval for the ECHILD project was granted by the National Research Ethics Service (17/LO/1494), NHS Health Research Authority Research Ethics Committee (20/EE/0180) and UCL Great Ormond Street Institute of Child Health’s Joint Research and Development Office (20PE06).

## Data Sharing and Access

Aggregate results from the ECHILD dataset will be pre-printed, revised and published. Individual record-level data with personal identifiers removed is currently hosted on the Office for National Statistics Secure Research Service’s data-sharing service. We are grateful to the Office for National Statistics (ONS) for providing the trusted research environment for the ECHILD Database. ONS agrees that the figures and descriptions of results in the attached document may be published. This does not imply ONS’ acceptance of the validity of the methods used to obtain these figures, or of any analysis of the results.

The ECHILD Database uses data from the Department for Education (DfE). The DfE does not accept responsibility for any inferences or conclusions derived by the authors. This work uses data provided by patients and collected by the National Health Service as part of their care and support. Source data can also be accessed by researchers by applying to NHS Digital.

## Appendix

**Appendix 1.**
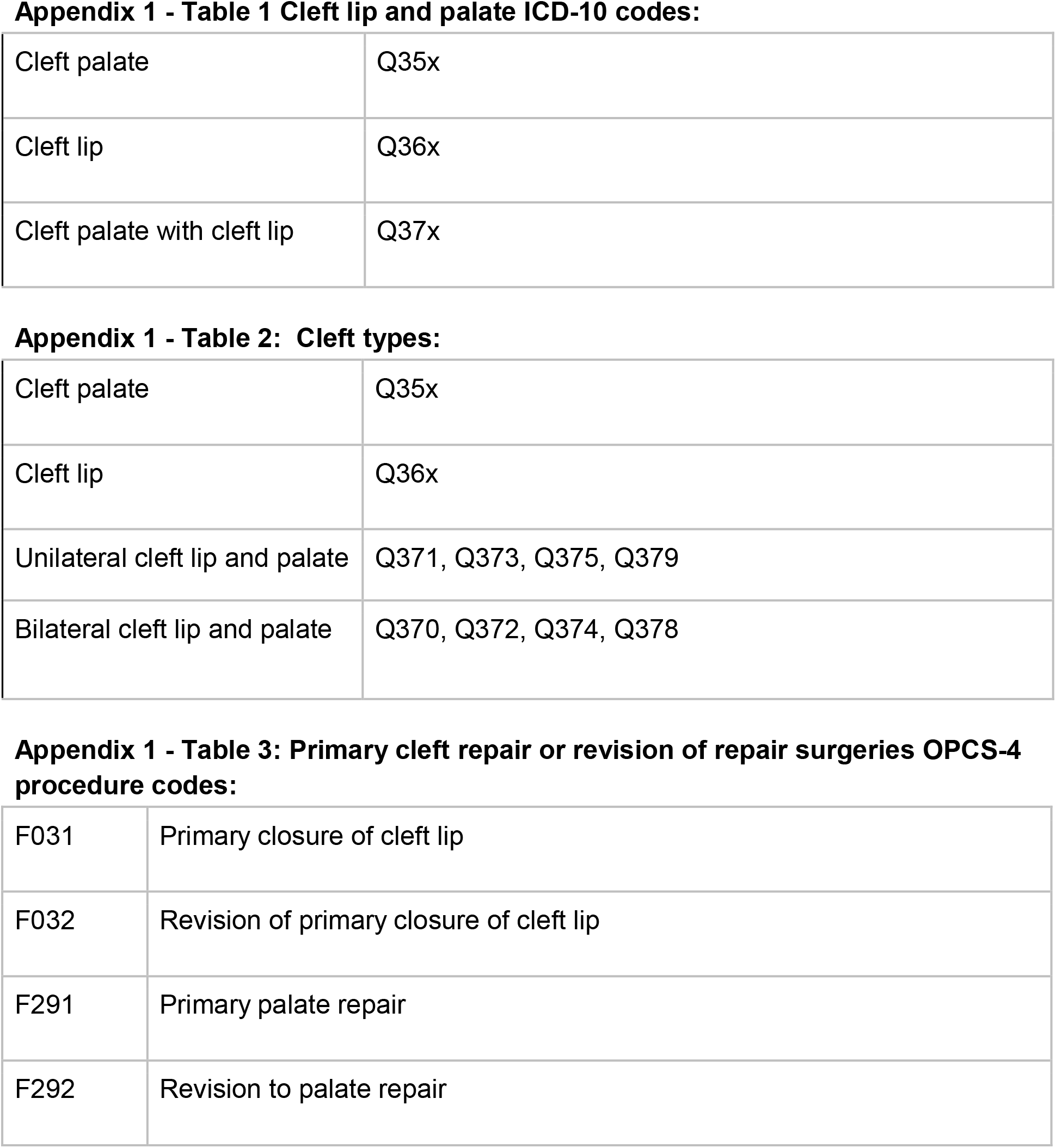
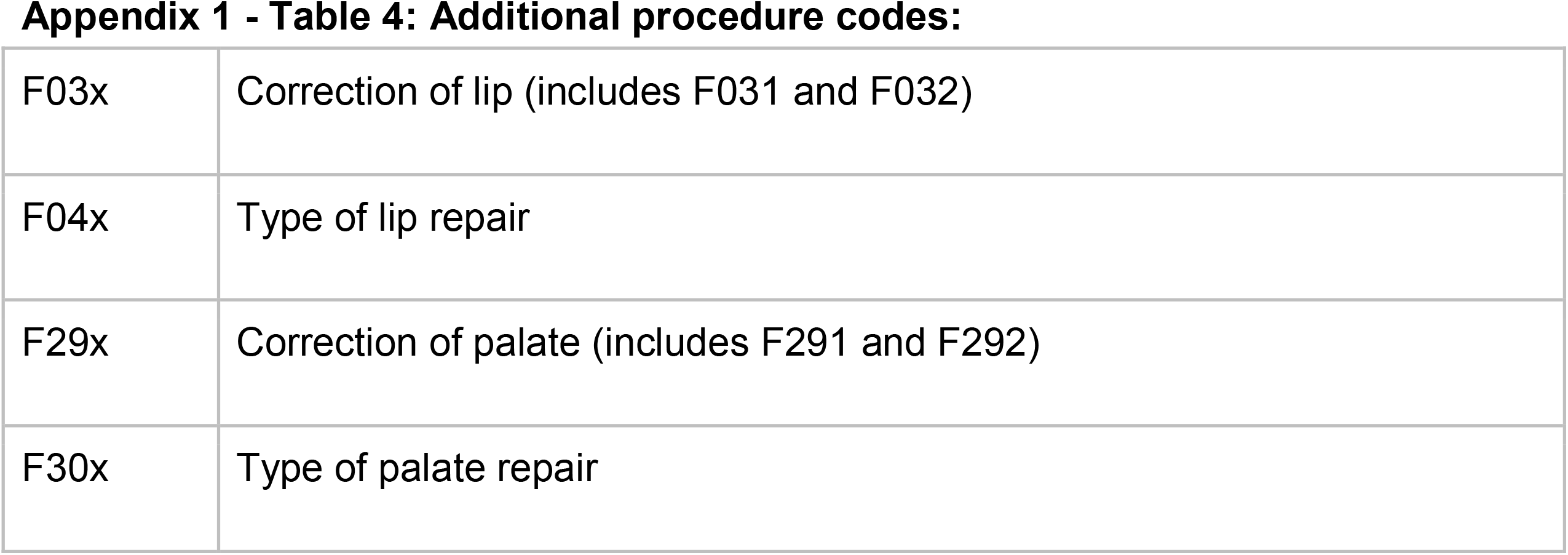
Phenotypes (ICD10 - OPCS) codes for cleft lip and palate.

|  |  |
| --- | --- |
| Cleft palate | Q35x |
| Cleft lip | Q36x |
| Cleft palate with cleft lip | Q37x |
| Appendix 1 - Table 2: Cleft types: |  |
| Cleft palate | Q35x |
| Cleft lip | Q36x |
| Unilateral cleft lip and palate | Q371, Q373, Q375, Q379 |
| Bilateral cleft lip and palate | Q370, Q372, Q374, Q378 |
| Appendix 1 - Table 3: Primary cleft repair or revision of repair surgeries OPCS-4 procedure codes: |  |
| F031 | Primary closure of cleft lip |
| F032 | Revision of primary closure of cleft lip |
| F291 | Primary palate repair |
| F292 | Revision to palate repair |
| **Appendix 1 - Table 4: Additional procedure codes:** |  |
| F03x | Correction of lip (includes F031 and F032) |
| F04x | Type of lip repair |
| F29x | Correction of palate (includes F291 and F292) |
| F30x | Type of palate repair |

**Appendix 2:** Determining unplanned admissions in hospital episode statistics admitted patient care.

| **Value of *admimeth* variable in HES** | **Meaning** | **Action** |
| --- | --- | --- |
| **11, 12, 13** | Planned admission | Planned |
| **21, 22, 23, 24, 25, 28, 2A, 2B, 2D** | Unplanned admission | Unplanned |
| **2C** | Unplanned admission for a baby born at home as intended (available from 2013/14) | Other (birth) |
| **31, 32** | Maternity admission | Other (maternity) |
| **82, 83** | Birth of a baby | Other (birth) |
| **81** | Transfer of any admitted patient from other Hospital Provider other than in an Unplanned | Planned |
| **84** | Admission by Admissions Panel of a High-Security Psychiatric Hospital, patient not entered on the HSPH Admissions Waiting List (available between 1999 and 2006) | Planned |
| **89** | HSPH Admissions Waiting List of a High-Security Psychiatric Hospital (available between 1999 and 2006) | Planned |
| **98** | Not applicable (available from 1996/97) | Other |
| **99** | Not known: a validation error | Other |

**Appendix 3:**
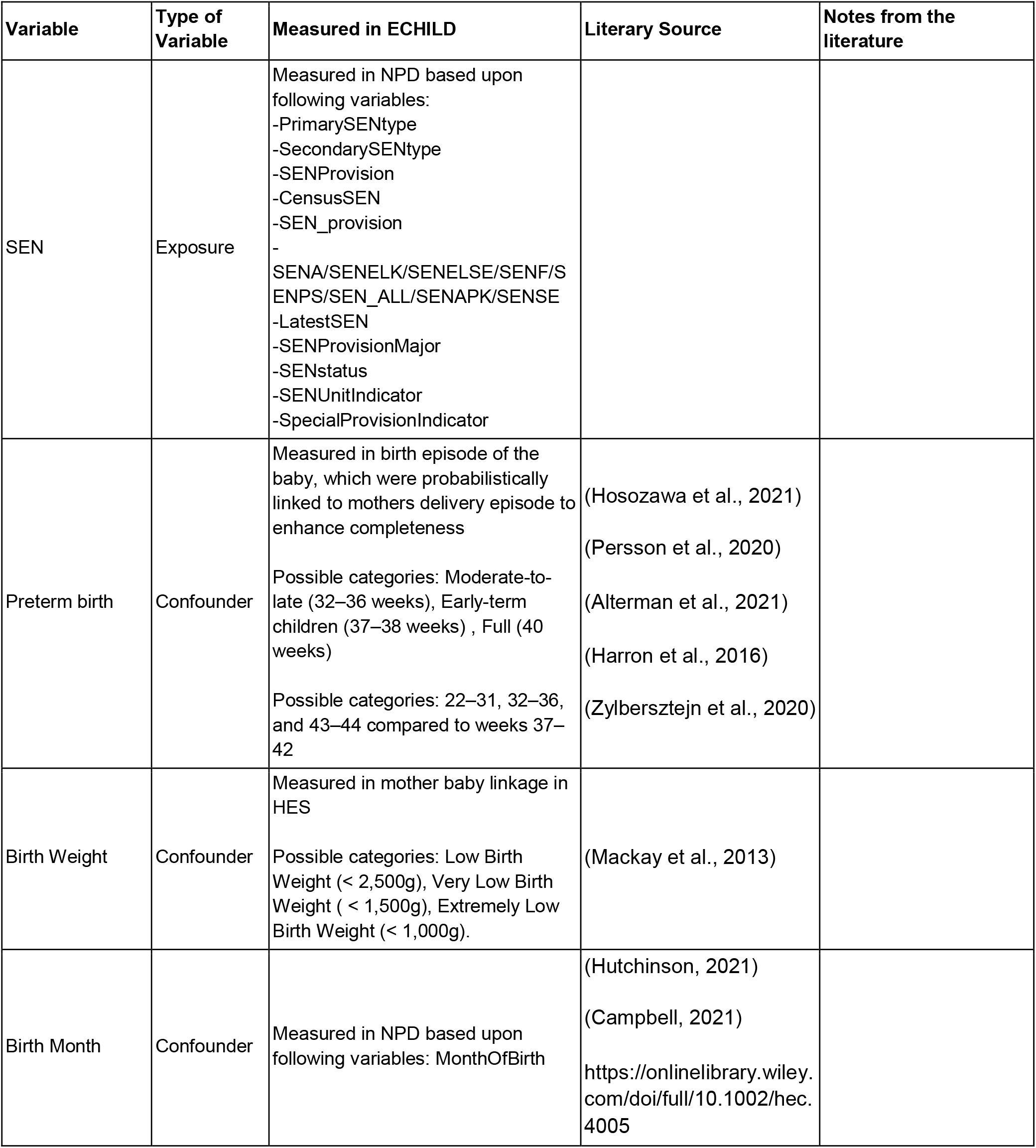
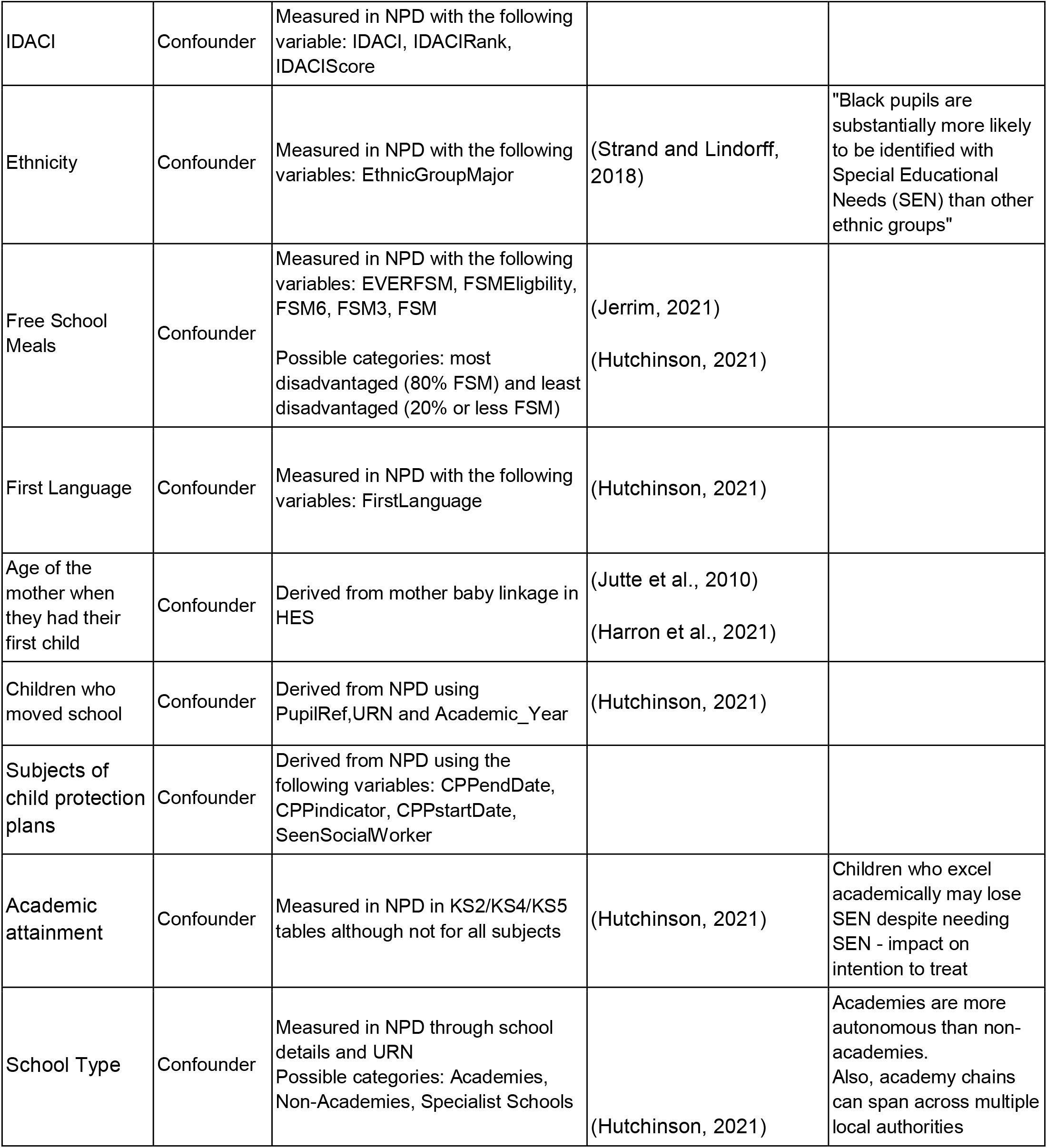
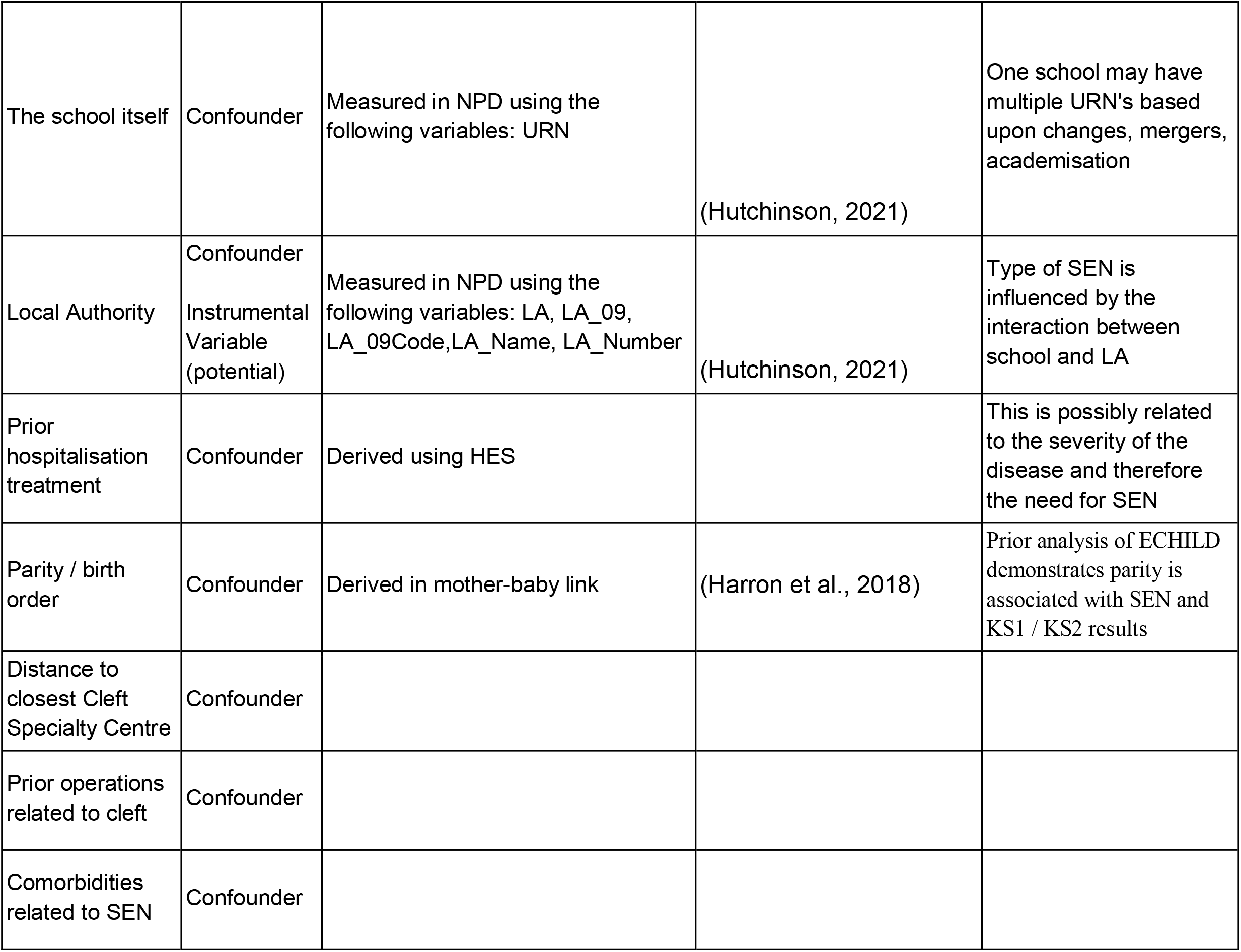
Variables related to receiving SEN and to unplanned hospitalisations.

**Appendix 4.**
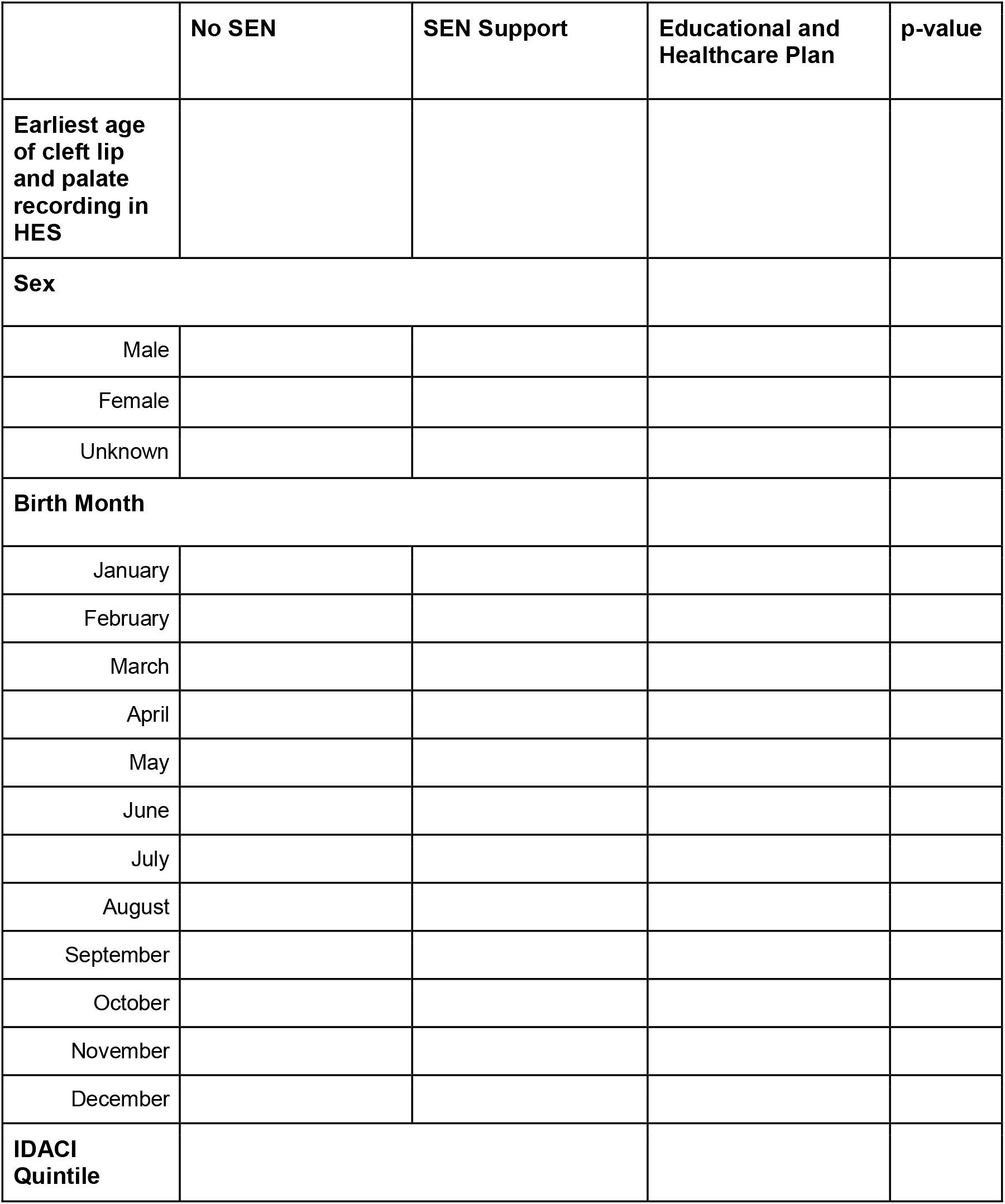
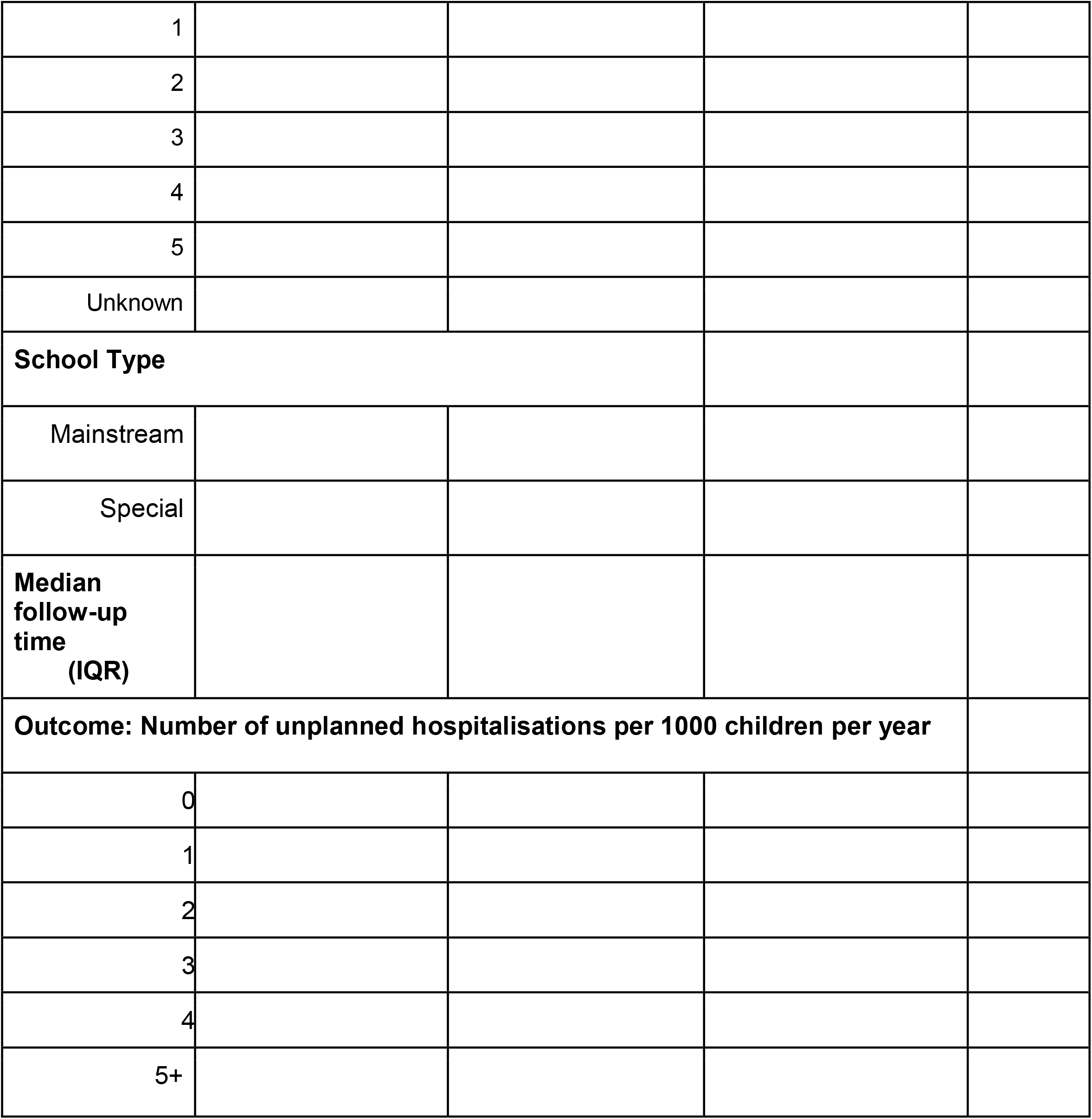
For each cleft severity level (Cleft Lip only, Cleft Palate only, Unilateral Cleft Lip and Palate, Bilateral Cleft Lip and Palate) and comorbidity (no comorbidities, at last one comorbidity) interaction, we will generate a separate distribution table. The table will include N and row percentages.

